# Initial Staging ^18^F-FDG PET/CT for Coronary Artery Calcium Scoring to Assess Cardiovascular Risk in Women with Breast Cancer

**DOI:** 10.64898/2026.08.24.26361275

**Authors:** Matthew R. Fleming, Kevin G. Tayon, Artur Schneider, Alyssa D. McPherson, Sidney M. Bianco, Ephraim E. Parent, Akarsh Sharma, Grace Lin, Nadine Norton, Jordan C. Ray

## Abstract

**Background.:** Cardiovascular disease is a leading cause of death among women with breast cancer, and the 2026 ACC/AHA dyslipidemia guideline endorses coronary artery calcium (CAC) scoring to guide statin therapy before cardiotoxic treatment. Breast cancer patients routinely undergo staging ^18^F-fluorodeoxyglucose PET/CT, whose low-dose CT visualizes the coronary arteries, thus enabling CAC quantification at no additional cost or radiation.

**Methods:** In this single-center retrospective study, consecutive women with newly diagnosed breast cancer undergoing staging ^18^F-FDG PET/CT (2009–2021) had semi-automated Agatston CAC scoring performed on the low-dose CT and were stratified by CAC presence (CAC-P) versus absence (CAC-A). We assessed a composite of cardiac diagnostic testing (stress testing, coronary CT angiography, invasive angiography), clinical events, and reclassification of statin eligibility per ACC/AHA guideline thresholds in a prevention-eligible subgroup.

**Results:** Among 276 women (mean age 55.5 years; median follow-up 7.1 years), CAC was present in 68 (25%) but was clinically reported in only 5.4%. CAC-P was associated with more cardiac testing (34% vs 12%; age-adjusted hazard ratio 2.75, 95% CI 1.43–5.28) and, though underpowered, with more atherosclerotic events (7.4% vs 1.4%), but not with the all-cause composite. In the prevention-eligible subgroup (n=39), CAC scoring would have changed statin eligibility in 64%, initiating therapy in 62% of CAC-P women and supporting de-prescribing in 67% of CAC-A women.

**Conclusions:** CAC can be feasibly quantified from staging PET/CT in women with breast cancer and would frequently reclassify statin eligibility at no additional cost or radiation, yet is rarely reported.

## Introduction

Cardiovascular disease (CVD) is a leading cause of morbidity and mortality among women with breast cancer, and among older and long-term survivors, particularly those with early-stage, hormone receptor–positive disease, CVD rivals or overtakes the cancer itself as a cause of death.^1,2^ This convergence reflects both shared risk factors and the cardiovascular toxicity of cancer therapy: anthracyclines, human epidermal growth factor receptor 2 (HER-2)–targeted agents, and thoracic radiotherapy each increase cardiovascular risk, and radiotherapy and chemotherapy have specifically been linked to accelerated atherosclerotic cardiovascular disease (ASCVD).^3,4^ Statins are a cornerstone of primary prevention, yet high-risk women are less likely than men to remain on guideline-directed statin therapy over time.^5^ Because cardiovascular risk rises once cardiotoxic treatment begins, the optimal time to characterize baseline risk and to act on it is at the time of cancer diagnosis, before therapy is initiated.

Coronary artery calcium (CAC) is among the most powerful tools for refining ASCVD risk and individualizing statin decisions, and its value is bidirectional.

Successive ACC/AHA guidelines have endorsed CAC scoring to individualize statin decisions in adults at borderline-to-intermediate risk. The current 2026 ACC/AHA dyslipidemia guideline recommends selective CAC scoring in women older than 45 years (and men older than 40) at borderline or intermediate risk and, restoring explicit lipid targets, supports an LDL-cholesterol goal below 100 mg/dL for anyone with detectable calcium, with lower targets as the score rises.^6^ The specific thresholds applied in this study derive from the 2019 ACC/AHA primary prevention guideline in force during the study period: in adults aged 40–75 years, a coronary artery calcium score (CACS) >100 Agatston units (AU) or ≥75th percentile favors therapy; 1–99 AU confers more modest benefit; and a score of 0 AU may justify withholding or deferring a statin.^7^ This “power of zero,” the ability to safely de-escalate therapy in a calcium-free artery, is particularly relevant to women with breast cancer, who carry high medication burdens and competing toxicities, where avoiding an unnecessary statin has tangible value.^8^ In oncology, however, guidance is contradictory. A 2022 Society of Cardiovascular Computed Tomography consensus recommends CAC assessment in cancer patients before therapy, whereas the 2022 European Society of Cardiology cardio-oncology guideline does not recommend routine CAC scoring prior to treatment but still advises reporting CAC when present on non-gated CT to guide preventive therapy.^9,10^ This unresolved tension has been highlighted as a gap in the evidence base.^11^ Its practical consequence is that, even when calcium is visible on imaging already performed, it is frequently neither scored nor reported.^12^

Breast cancer patients routinely undergo ^18^F-fluorodeoxyglucose positron emission tomography/computed tomography (^18^F-FDG PET/CT) for initial staging, and the low-dose CT component visualizes the coronary arteries. In keeping with the As Low As Reasonably Achievable principle, deriving CACS from these existing scans could spare patients the radiation and cost of a dedicated study. CAC measured on non-cardiac CT correlates closely with dedicated ECG-gated scoring and carries comparable prognostic weight,^13,14^ and prior work has scored CAC from PET/CT in predominantly mixed-cancer cohorts, using visual or ordinal grading,^15,16^ fully automated artificial intelligence,^17^ and, recently, to reclassify primary-prevention management.^18^ In breast cancer specifically, CAC has been studied chiefly on radiotherapy-planning CT,^19,20^ which is acquired at treatment and excludes patients who do not receive radiotherapy. What remains unestablished is whether a discrete, percentile-capable Agatston score, the value required to apply the full set of guideline thresholds, including the 75th-percentile statin trigger, can be obtained semi-automatically from the earliest, pre-treatment staging PET/CT in an all-female breast-cancer population; how often doing so would change guideline-based statin eligibility in either direction; and how frequently CAC is captured during routine clinical interpretation.

We therefore studied women with newly diagnosed breast cancer to address four objectives: 1) to determine the feasibility of semi-automatic Agatston CAC quantification from the low-dose CT of initial staging ^18^F-FDG PET/CT; 2) to estimate how often a CACS derived in this way would reclassify guideline-based statin eligibility, both initiating therapy in under-treated patients and supporting de-prescribing when CAC is absent; 3) to characterize the association of CAC with downstream cardiac testing and clinical events; and 4) to quantify how often CAC was reported during routine clinical PET/CT interpretation. To our knowledge, this is the first study to address these questions in an all-female breast-cancer cohort using pre-treatment staging PET/CT.

## Methods

### Data Availability

The data that support the findings of this study are available from the corresponding author upon reasonable request. The data are not publicly available because they contain protected health information and are subject to institutional restrictions.

### Study Design and Population

This was a single-center retrospective cohort study of consecutive women with newly diagnosed breast cancer referred for initial staging ^18^F-FDG PET/CT between February 1, 2009, and July 31, 2021. Patients with uninterpretable images or incomplete data sets were excluded. The study was approved by the Mayo Clinic Institutional Review Board (IRB 22-013359), which granted a waiver of informed consent, and was conducted in accordance with the Declaration of Helsinki. Reporting follows the STROBE guideline for observational studies.

Patient demographics, medical and cardiovascular history, breast cancer grade, receptor status (estrogen receptor, progesterone receptor, HER-2), serum lipids within one year of the PET/CT, and blood pressure on the day of the scan were obtained from the electronic medical record (Epic, Epic Systems Corporation, Verona, WI). ASCVD risk scores, Multi-Ethnic Study of Atherosclerosis (MESA) scores, and arterial age were calculated using data available at the time of the PET/CT.^21,22,23^

### Image Acquisition and CAC Scoring

PET/CT studies were performed in standard clinical practice. Scanner platforms and acquisition parameters evolved over the study period; representative current institutional protocols use non-gated acquisition at 100–140 kVp with automated tube-current modulation and a reconstructed slice thickness of 3.0–4.0 mm. The low-dose, non-contrast CT was reviewed in axial reconstruction with windowing optimized for calcium. CAC was identified by its location within the coronary arteries and attenuation >130 Hounsfield units (HU), and a semi-automatic Agatston score was computed^24^ using commercially available calcium-scoring software. Scores were reported in Agatston units and as a percentile for age, sex, and race.^25^ Equivocal findings were resolved by consensus.

### Outcomes

The primary outcome was a composite of all-cause death, non-fatal myocardial infarction (Fourth Universal Definition^26^), and coronary revascularization (percutaneous intervention or bypass). The secondary outcome was a composite of cardiac diagnostic testing: stress testing, coronary CT angiography (CCTA), and invasive coronary angiography. In a post hoc analysis, major adverse cardiac events (MACE) were defined as a composite of cardiovascular death, non-fatal myocardial infarction, and coronary revascularization; this composite comprised atherosclerotic events only and did not include heart failure, stroke, or cancer therapy–related cardiac dysfunction. Only events occurring after the staging PET/CT were counted. Follow-up was censored administratively on July 31, 2023.

### Statin-Eligibility Subgroup

A prespecified subgroup comprised women aged 40–75 years with borderline (5 to <7.5%) or intermediate (7.5 to <20%) ASCVD risk and LDL <190 mg/dL. To isolate patients for whom CAC would guide a primary-prevention decision, those with an independent statin indication (diabetes mellitus, established ASCVD, or LDL ≥190 mg/dL) were excluded (n=39; a sensitivity analysis retaining diabetes and established ASCVD comprised n=57). The 2019 ACC/AHA thresholds (CACS >100 AU, ≥75th percentile, or age >55 years with CACS 1–99 AU) were applied to determine whether CAC scoring would change statin eligibility in either direction.

### Statistical Analysis

Continuous variables are presented as mean ± SD and compared with the t-test; categorical variables as frequencies and compared with the Fisher exact or chi-square test. Coronary artery calcium was analyzed both dichotomously (present vs absent) and across ordinal categories (0, 1–99, 100–399, and ≥400 AU), with a Cox model used to test the trend across categories. Time-to-event analyses used Kaplan-Meier estimates with the log-rank test and Cox proportional-hazards models, reported unadjusted and adjusted for age (primary analysis); models additionally adjusted for the ASCVD risk score were performed as a sensitivity analysis among patients with a calculable score. The proportional-hazards assumption was assessed with Schoenfeld residuals, and effect modification by age was tested with a multiplicative interaction term. Because non-cardiovascular death is a competing event, cumulative incidence functions for major adverse cardiac events and for cardiac testing were additionally estimated using the Aalen-Johansen method (treating death as a competing risk) and compared between groups with Gray’s test. ASCVD risk scores were calculated with the Pooled Cohort Equations and MESA scores with the MESA risk calculator where the requisite data were available; all analyses were complete-case, and a complete lipid profile, required for these scores, was available in the medical record for 174 patients (63%).

Reclassification of statin eligibility was analyzed descriptively among patients with a calculable ASCVD risk score. A two-sided p<0.05 was considered significant. Analyses were performed in Python 3 (SciPy, lifelines, statsmodels) and GraphPad Prism. Dr Fleming had full access to all the data in the study and takes responsibility for its integrity and the data analysis.

## Results

### Cohort and feasibility of CAC scoring

A semi-automatic Agatston score was successfully obtained from the low-dose CT of the staging ^18^F-FDG PET/CT in all 276 women (mean age 55.5 ± 11.8 years; 100% female). Over a median follow-up of 7.1 years (reverse Kaplan-Meier), coronary artery calcium was present (CAC-P) in 68 women (25%) and absent (CAC-A) in 208 (75%). All participants were women; self-reported race or ethnicity was White in 241 (87.3%), Black in 28 (10.1%), Hispanic in 5 (1.8%), and Asian in 2 (0.7%). Because the cohort was exclusively female and predominantly White, sex-based comparisons were not possible and the study was not powered to detect race- or ethnicity-based differences in outcomes. Among CAC-P women, the median CACS was 68.2 AU (mean 218.2 ± 556.8 AU), corresponding to the 74th percentile for age, sex, and race (available for 61 of 68 CAC-P women). As shown in Table 1, CAC-P women were older (63.9 vs 52.7 years, p<0.001) and had higher ASCVD risk scores (10.8% vs 4.6%, p<0.001), but the groups did not differ in body-mass index, systolic blood pressure, LDL, prevalence of traditional risk factors, tumor grade, or hormone-receptor status. Despite this calcium burden, CAC was noted in the original clinical PET/CT report in only 15 of 276 women (5.4%), and reporting did not increase with calcium burden; calcium was documented in none of the six women with a score ≥400 AU (Table 2).

**Table 1.** Baseline characteristics.

| Baseline characteristic | Total (n=276) | CAC-P (n=68) | CAC-A (n=208) | p |
| --- | --- | --- | --- | --- |
| Age, years | 55.5 ± 11.8 | 63.9 ± 11.4 | 52.7 ± 10.6 | <0.001 |
| ASCVD 10-yr risk, %* | 6.2 ± 7.5 | 10.8 ± 9.4 | 4.6 ± 5.9 | <0.001 |
| BMI, kg/m<sup>2</sup> | 28.3 ± 6.5 | 28.2 ± 5.9 | 28.4 ± 6.7 | 0.87 |
| SBP, mmHg | 127.7 ± 18.4 | 127.6 ± 18.2 | 127.8 ± 18.5 | 0.93 |
| LDL, mg/dL | 107.7 ± 33.0 | 114.0 ± 36.8 | 105.5 ± 31.4 | 0.14 |
| Breast cancer grade | 2.5 ± 0.7 | 2.4 ± 0.7 | 2.5 ± 0.7 | 0.74 |
| History of CAD | 8/276 (3%) | 2/68 (3%) | 6/208 (3%) | 1.00 |
| Hyperlipidemia | 114/276 (41%) | 23/68 (34%) | 91/208 (44%) | 0.16 |
| Hypertension | 122/276 (44%) | 25/68 (37%) | 97/208 (47%) | 0.16 |
| Diabetes mellitus | 41/275 (15%) | 9/68 (13%) | 32/207 (15%) | 0.85 |
| Smoking history | 17/276 (6%) | 7/68 (10%) | 10/208 (5%) | 0.14 |
| Family history of CAD | 32/276 (12%) | 7/68 (10%) | 25/208 (12%) | 0.83 |
| ER positive | 192/275 (70%) | 44/68 (65%) | 148/207 (71%) | 0.29 |
| PR positive | 163/275 (59%) | 40/68 (59%) | 123/207 (59%) | 1.00 |
| HER-2 positive | 78/272 (29%) | 22/68 (32%) | 56/204 (27%) | 0.44 |
| Coronary artery calcium score, AU | 53.8 ± 290.5 | 218.2 ± 556.8 | 0 ± 0 | <0.001 |

Continuous values are mean ± SD; categorical values are n/N (%). Denominators vary
where individual data points were unavailable. \*ASCVD risk score calculable in 174 (63%) with a complete lipid profile. ASCVD indicates atherosclerotic cardiovascular disease; BMI, body mass index; CAC-A, coronary artery calcium absent; CAC-P, coronary artery calcium present; CAD, coronary artery disease; ER, estrogen receptor; HER-2, human epidermal growth factor receptor 2; LDL, low-density lipoprotein; PR, progesterone receptor; and SBP, systolic blood pressure.

**Table 2.**
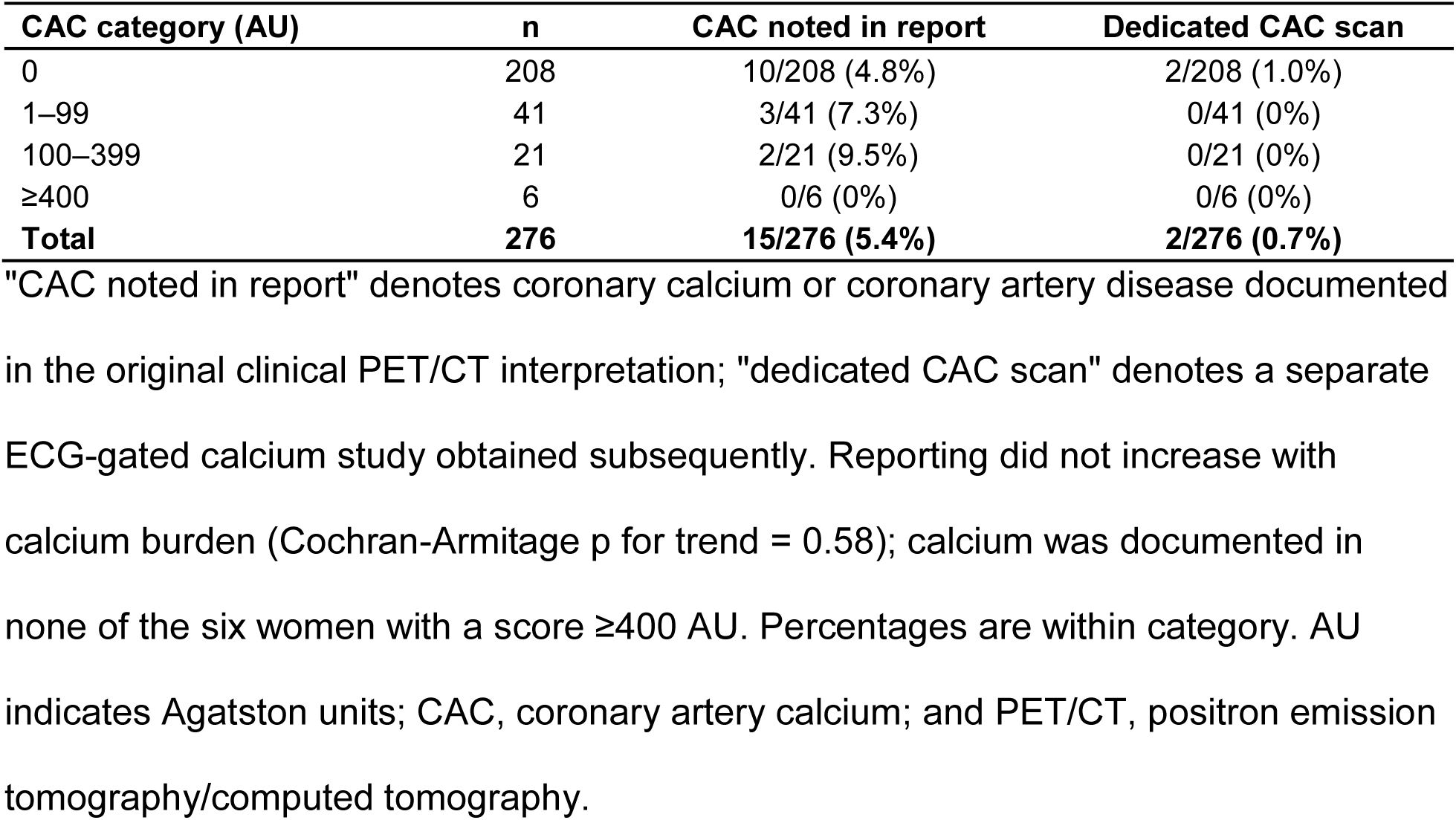
Clinical reporting of coronary artery calcium by calcium burden.

| **CAC category (AU)** | **n** | **CAC noted in report** | **Dedicated CAC scan** |
| --- | --- | --- | --- |
| 0 | 208 | 10/208 (4.8%) | 2/208 (1.0%) |
| 1–99 | 41 | 3/41 (7.3%) | 0/41 (0%) |
| 100–399 | 21 | 2/21 (9.5%) | 0/21 (0%) |
| ≥400 | 6 | 0/6 (0%) | 0/6 (0%) |
| **Total** | **276** | **15/276 (5.4%)** | **2/276 (0.7%)** |

"CAC noted in report" denotes coronary calcium or coronary artery disease documented in the original clinical PET/CT interpretation; "dedicated CAC scan" denotes a separate ECG-gated calcium study obtained subsequently. Reporting did not increase with calcium burden (Cochran-Armitage p for trend = 0.58); calcium was documented in none of the six women with a score ≥400 AU. Percentages are within category. AU indicates Agatston units; CAC, coronary artery calcium; and PET/CT, positron emission tomography/computed tomography.

### CAC and reclassification of statin eligibility

Among the 39 women eligible for CAC- guided primary prevention, applying the 2019 ACC/AHA thresholds to the PET/CT-derived CACS would have changed management in 25 (64%). The effect was bidirectional (Table 3): among 21 CAC-P women, 13 (62%) were statin-eligible yet untreated and would newly qualify for therapy, whereas among 18 CAC-A women, 12 (67%) were receiving a statin without a guideline indication and could be considered for de-prescribing. Within the CAC-P group, 7 of 21 had a CACS ≥100 AU, 7 had a CACS ≥75th percentile, and 14 met the age >55 years with CACS 1–99 AU criterion. Results were consistent in a sensitivity analysis that also included women with diabetes or established ASCVD (n=57; management changed in 30 [53%]).

**Table 3.** Statin-eligibility subgroup.

| **Secondary analysis** | **Total (n=39)** | **CAC-P (n=21)** | **CAC-A (n=18)** | **p** |
| --- | --- | --- | --- | --- |
| CACS, AU | 41.8 ± 75.3 | 77.7 ± 88.6 | 0 ± 0 | <0.001 |
| Borderline ASCVD risk | 8/39 (21%) | 5/21 (24%) | 3/18 (17%) | — |
| Intermediate ASCVD risk | 31/39 (79%) | 16/21 (76%) | 15/18 (83%) | — |
| CACS ≥100 AU | 7/39 (18%) | 7/21 (33%) | 0/18 (0%) | 0.008 |
| CACS ≥75th percentile | 7/39 (18%) | 7/21 (33%) | 0/18 (0%) | 0.008 |
| CACS 1–99 AU and age >55 | 14/39 (36%) | 14/21 (67%) | 0/18 (0%) | <0.001 |
| CACS changes management | 25/39 (64%) | 13/21 (62%) | 12/18 (67%) | 0.75 |
| Not on statin & indicated | 13/39 (33%) | 13/21 (62%) | 0/18 (0%) | <0.001 |
| On statin & not indicated | 12/39 (31%) | 0/21 (0%) | 12/18 (67%) | <0.001 |
| LDL, mg/dL | 111 ± 35 | 123 ± 36 | 97 ± 28 | 0.017 |

Primary analysis, n=39; patients with diabetes, established ASCVD, or LDL ≥190 mg/dL
were excluded. ASCVD indicates atherosclerotic cardiovascular disease; AU, Agatston units; CAC-A, coronary artery calcium absent; CAC-P, coronary artery calcium present; CACS, coronary artery calcium score; and LDL, low-density lipoprotein.

### Graded relationship with calcium burden

Associations strengthened progressively with increasing calcium burden (Table 4). Compared with a CACS of 0, event rates rose across the 1–99, 100–399, and ≥400 AU categories for cardiac testing (12%, 32%, 38%, 33%; p for trend = 0.0003), major adverse cardiac events (1.4%, 4.9%, 9.5%, 16.7%; p for trend = 0.006), and, less consistently, all-cause death (17%, 10%, 29%, 67%; p for trend = 0.04), the mortality trend being driven largely by the highest category, which included only six women.

**Table 4.** Outcomes across coronary artery calcium categories.

| **CAC category (AU)** | **n** | **Cardiac testing** | **MACE** | **All-cause death** |
| --- | --- | --- | --- | --- |
| 0 | 208 | 25/208 (12%) | 3/208 (1.4%) | 36/208 (17%) |
| 1–99 | 41 | 13/41 (32%) | 2/41 (4.9%) | 4/41 (10%) |
| 100–399 | 21 | 8/21 (38%) | 2/21 (9.5%) | 6/21 (29%) |
| ≥400 | 6 | 2/6 (33%) | 1/6 (16.7%) | 4/6 (67%) |
| **p for trend** |  | **0.0003** | **0.006** | **0.04** |

p for trend from a Cox model across ordinal categories. AU indicates Agatston units;
CAC, coronary artery calcium; and MACE, major adverse cardiac events.

### CAC and cardiac testing utilization

Women with CAC subsequently underwent more cardiac diagnostic testing, which occurred in 34% versus 12% over follow-up (cause-specific HR 2.91, 95% CI 1.64–5.16, log-rank p<0.001), an association that persisted after adjustment for age (HR 2.75, 95% CI 1.43–5.28, p=0.002). Accounting for the competing risk of death, the 10-year cumulative incidence of testing was 34% versus 15% (Gray’s test p<0.001; Figure 1). As shown in Table 5, testing was driven by stress testing (28% vs 11%, p=0.001) and invasive coronary angiography (8.8% vs 1.0%, p=0.003), with a non-significant trend toward more coronary CT angiography (5.9% vs 1.4%, p=0.065). The association was attenuated and no longer significant after additional adjustment for the ASCVD risk score in the subset with a calculable score (HR 1.97, 95% CI 0.87–4.42), a comparison limited by the smaller number of patients and events. This utilization endpoint is descriptive and hypothesis-generating.

**Figure 1.**
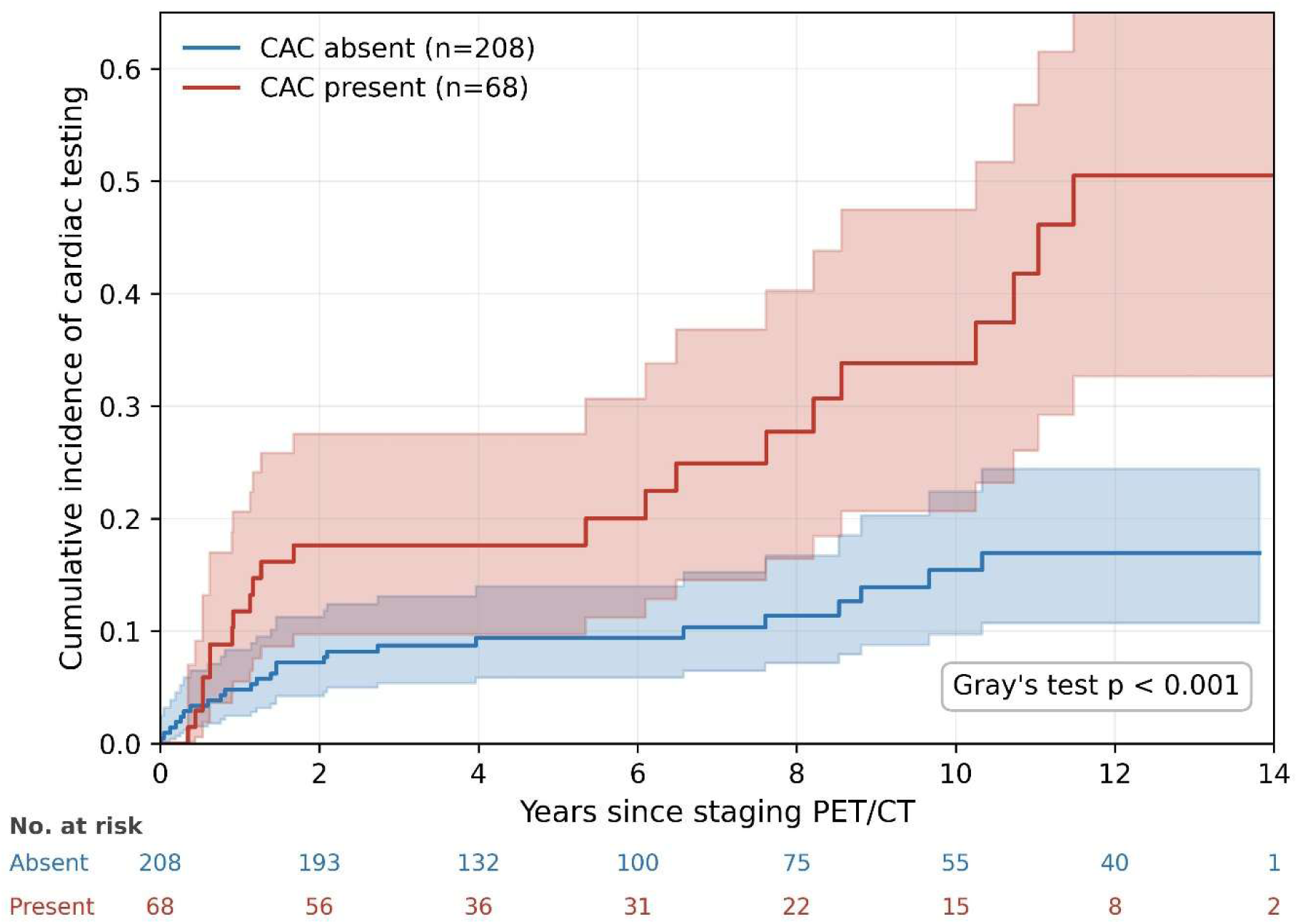
Cumulative incidence of downstream cardiac diagnostic testing according to coronary artery calcium status. Cardiac testing comprised stress testing, coronary computed tomography angiography, and invasive coronary angiography. Cumulative incidence was estimated with the Aalen-Johansen method, treating death as a competing risk, and compared between groups with Gray’s test. Women with coronary artery calcium underwent substantially more testing over follow- up (10-year cumulative incidence 34% versus 15%; p < 0.001). Shaded bands denote 95% confidence intervals and numbers at risk are shown below the x-axis. This utilization endpoint is descriptive and hypothesis-generating. CAC indicates coronary artery calcium; and PET/CT, positron emission tomography/computed tomography.

**Table 5.** Clinical endpoints and risk-reclassification metrics.

| Outcome / measure | Total (n=276) | CAC-P (n=68) | CAC-A (n=208) | p |
| --- | --- | --- | --- | --- |
| Primary composite outcome | 55/276 (20%) | 17/68 (25%) | 38/208 (18%) | 0.23 |
| All-cause death | 50/276 (18%) | 14/68 (21%) | 36/208 (17%) | 0.59 |
| Cardiovascular death | 1/276 (0.4%) | 0/68 (0%) | 1/208 (0.5%) | 1.00 |
| Non-fatal MI | 6/276 (2.2%) | 4/68 (5.9%) | 2/208 (1.0%) | 0.034 |
| Coronary revascularization | 1/276 (0.4%) | 1/68 (1.5%) | 0/208 (0%) | 0.25 |
| Major adverse cardiac events (MACE)<sup>‡</sup> | 8/276 (2.9%) | 5/68 (7.4%) | 3/208 (1.4%) | 0.024 |
| Secondary composite outcome | 48/276 (17%) | 23/68 (34%) | 25/208 (12%) | <0.001 |
| Stress testing | 41/276 (15%) | 19/68 (28%) | 22/208 (11%) | 0.001 |
| CCTA | 7/276 (2.5%) | 4/68 (5.9%) | 3/208 (1.4%) | 0.065 |
| Invasive coronary angiography | 8/276 (2.9%) | 6/68 (8.8%) | 2/208 (1.0%) | 0.003 |
| MESA score without CACS | 3.8 ± 3.4 | 5.1 ± 4.4 | 3.3 ± 2.8 | 0.004 |
| MESA score with CACS<sup>†</sup> | 3.1 ± 3.5 | 6.4 ± 5.1 | 1.7 ± 0.9 | <0.001 |
| Arterial age, years<sup>*</sup> | 46.2 ± 14.0 | 68.3 ± 12.0 | 39.0 ± 0.0 | — |

p values are from Fisher exact or chi-square tests for proportions; time-to-event
comparisons (log-rank/Cox) are reported in the text. <sup>\*</sup>Arterial age is set to a floor of 39 years when CACS = 0 (MESA formula); the between-group comparison is therefore not statistically meaningful and no p value is reported. <sup>†</sup>MESA scores calculable in 143 (52%) with a complete lipid profile. <sup>‡</sup>Composite of non-fatal MI, cardiovascular death, and coronary revascularization. CAC-A indicates coronary artery calcium absent; CAC-P, coronary artery calcium present; CACS, coronary artery calcium score; CCTA, coronary computed tomography angiography; MACE, major adverse cardiac events; MESA, Multi-Ethnic Study of Atherosclerosis; and MI, myocardial infarction.

### CAC and clinical events

Overall survival did not differ by CAC status: all-cause mortality was similar in women with and without calcium (21% vs 17%; HR 1.13, 95% CI 0.61–2.10; log-rank p=0.69; Figure 2) and remained non-significant after adjustment for age (HR 0.99, 95% CI 0.50–1.97). The primary composite outcome (all-cause death, non-fatal myocardial infarction, coronary revascularization) likewise did not differ between groups (17/68 [25%] vs 38/208 [18%]; HR 1.36, 95% CI 0.77–2.40, p=0.30; age-adjusted HR 1.15, 95% CI 0.60–2.19). Because mortality in this cohort was overwhelmingly non-cardiovascular (only one cardiovascular death occurred), we additionally examined a composite of major adverse cardiac events (MACE; non-fatal myocardial infarction, cardiovascular death, and coronary revascularization), which occurred more frequently in CAC-P women (5/68 [7.4%] vs 3/208 [1.4%]; cause-specific HR 4.95, 95% CI 1.18–20.7; log-rank p=0.015; Figure 3), although with only eight events the estimate was imprecise and no longer significant after age adjustment (HR 3.51, 95% CI 0.67–18.3). Accounting for competing non-cardiovascular deaths, the 10- year cumulative incidence of MACE was 5.8% versus 3.1%. Non-fatal myocardial infarction alone was also more common in CAC-P women (5.9% vs 1.0%, p=0.034).

**Figure 2.**
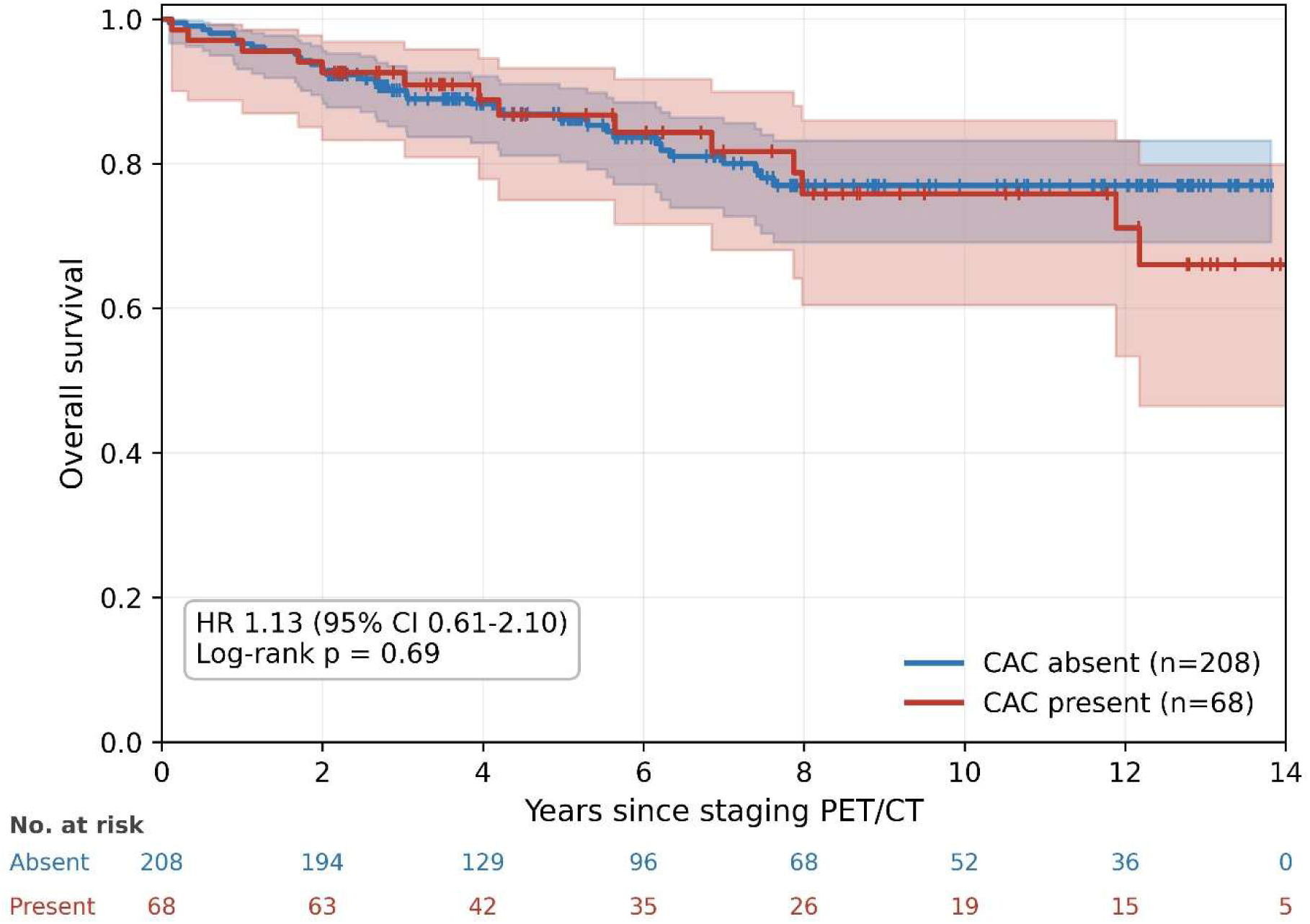
Overall survival according to coronary artery calcium status. Kaplan-Meier estimates of freedom from all-cause death are shown for women with and without coronary artery calcium identified on the low-dose computed tomography of staging ^18^F- FDG positron emission tomography/computed tomography. Survival did not differ between groups (hazard ratio 1.13, 95% CI 0.61–2.10; log-rank p = 0.69) and remained non-significant after adjustment for age. Mortality in this cohort was overwhelmingly non-cardiovascular; the primary composite outcome is reported in the text and Table 5. Shaded bands denote 95% confidence intervals and numbers at risk are shown below the x-axis. Follow-up was censored administratively on July 31, 2023. CAC indicates coronary artery calcium; CI, confidence interval; and ^18^F-FDG, ^18^F-fluorodeoxyglucose.

**Figure 3.**
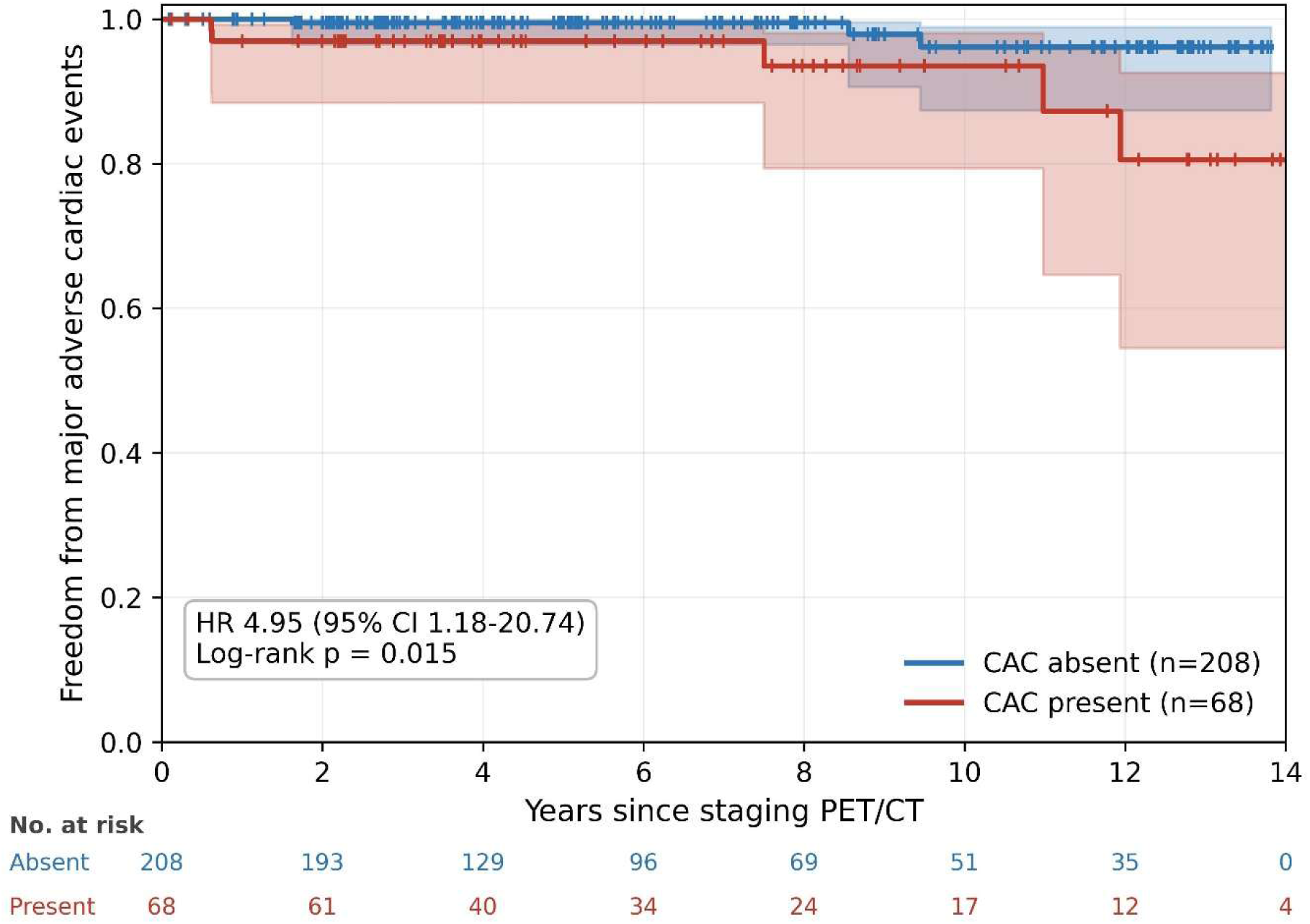
Freedom from major adverse cardiac events according to coronary artery calcium status. Major adverse cardiac events comprised cardiovascular death, non-fatal myocardial infarction, and coronary revascularization. Deaths without a preceding cardiac event were censored at the time of death (cause-specific analysis). Events occurred more frequently among women with coronary artery calcium (7.4% versus 1.4%; cause-specific hazard ratio 4.95, 95% CI 1.18–20.7; log-rank p = 0.015), although with only eight events the estimate is imprecise, as reflected in the wide confidence interval, and it did not remain significant after adjustment for age. Stroke, heart failure, and cancer therapy–related cardiac dysfunction were not captured. Shaded bands denote 95% confidence intervals and numbers at risk are shown below the x-axis. Follow-up was censored administratively on July 31, 2023. CAC indicates coronary artery calcium; CI, confidence interval; and MACE, major adverse cardiac events.

### Risk reclassification metrics

Incorporating the PET/CT-derived CACS into the MESA score reclassified estimated risk in both directions (Table 5), and in CAC-P women, the CAC-derived arterial age exceeded chronological age (68.3 vs 63.9 years, p=0.011).

### Model diagnostics

The proportional-hazards assumption was satisfied for all Cox models (Schoenfeld p ≥ 0.18). There was no significant effect modification by age for cardiac testing or major adverse cardiac events (interaction p = 0.74 and 0.15, respectively); a nominal interaction was observed for all-cause mortality (p = 0.03), which should be interpreted with caution given multiple testing.

## Discussion

In this single-center cohort of 276 women undergoing staging ^18^F-FDG PET/CT for newly diagnosed breast cancer, we report three principal findings. First, a discrete, percentile-capable Agatston coronary artery calcium score could be obtained semi-automatically from the low-dose CT in nearly all women. Second, applying guideline thresholds to that score would have reclassified statin eligibility in most prevention- eligible women, initiating therapy in the under-treated and supporting de-prescribing when calcium was absent. Third, despite a 25% prevalence of coronary calcium, it was documented in only 5.4% of clinical PET/CT reports. To our knowledge, this is the first study to quantify these effects in an all-female breast-cancer cohort using the earliest, pre-treatment imaging.

Although CAC is most often quantified from dedicated ECG-gated CT, calcium on non-gated and non-cardiac CT correlates closely with gated scoring and carries comparable prognostic weight.^13,14^ Prior efforts to score CAC from PET/CT have relied on visual or ordinal grading, which yields a category rather than a value,^15,16^ or on fully automated artificial intelligence.^17^ Our use of semi-automatic quantification is distinct in producing a discrete Agatston score, which, unlike an ordinal range, can be converted to the age-, sex-, and race-specific percentile that guidelines use to guide statin therapy, including the ≥75th-percentile threshold.^7,25^ Deriving this value from a scan already performed for cancer staging spares the patient the radiation (estimated 0.74–1.26 mSv) and cost (estimated $75–150) of a dedicated study,^27,28^ a meaningful consideration in a population already burdened by extensive oncologic imaging and expense.

The clearest clinical implication of our findings is therapeutic. Among prevention- eligible women, CAC scoring would have changed statin management in nearly two-thirds, with effects in both directions. On the one hand, the majority of women with calcium were statin-eligible yet untreated, echoing the well-documented underuse of guideline-directed statins in women.^5^ On the other hand, two-thirds of women without calcium were taking a statin despite lacking a guideline indication, a group for whom a calcium score of zero, the so-called “power of zero,” provides a rationale to de-escalate therapy.^8^ The value of de-prescribing is amplified in breast-cancer survivors, who take substantially more medications than their peers and face competing toxicities and drug interactions, so avoiding an unnecessary daily therapy carries a tangible benefit.^29,30^ A recent study reported that CAC on FDG-PET/CT reclassified primary-prevention management in a mixed-cancer population;^18^ our findings extend this specifically to women with breast cancer at the pre-treatment baseline and, importantly, document both the de-prescribing opportunity and statin initiation. These decisions remain aligned with current guidance: the 2026 ACC/AHA dyslipidemia guideline continues to endorse CAC-guided statin allocation in women at borderline-to-intermediate risk and specifically recommends continuing lipid-lowering therapy in patients undergoing cancer treatment, making a baseline calcium score obtained at staging directly actionable.^6^

That calcium was present in one in four women yet reported in only one in twenty exposes a substantial, correctable gap between what these scans contain and what reaches the treating clinician. This mirrors the broad under-reporting of incidental CAC on non-gated CT, present in reports only 19–93% of the time,^14^ and it likely reflects the discordant guidance clinicians face: a Society of Cardiovascular Computed Tomography consensus recommends CAC assessment before cancer therapy,^9^ whereas the European Society of Cardiology cardio-oncology guideline does not endorse routine scoring yet still advises reporting calcium when it is seen, a tension recently highlighted as a gap in the evidence.^10,11^ Documentation, moreover, did not reliably translate into action: among the 15 women in whom calcium was reported, a downstream change in preventive management was recorded in only one, though responses to the report were inconsistently captured and this figure should be interpreted with caution. Interventions as simple as templated reporting have been shown to increase appropriate statin initiation,^12^ and a baseline calcium score obtained at staging could also serve as the reference for the serial imaging increasingly recommended for cancer survivors.^31^

CAC was not associated with our primary composite clinical outcome, and only a single cardiovascular death occurred in the entire cohort. Rather than a null result to be explained away, this is the expected pattern in a relatively young, all-female population with a modest calcium burden. In the Coronary Artery Calcium Consortium, cancer is the leading cause of death when calcium is absent, and cardiovascular mortality overtakes cancer only at high calcium scores, thresholds that are higher in women and younger patients.^32^ Our cohort, with a median CAC score of 68 AU among affected women and mortality driven almost entirely by non-cardiovascular causes, is consistent with this framework and with a prior breast-cancer study in which CAC-associated deaths were overwhelmingly cancer-related.^19^ Notably, when mortality was removed and a cardiovascular-specific composite was examined, calcium was associated with a markedly higher event rate (7.4% vs 1.4%), suggesting the all-cause composite masked a cardiovascular signal. This estimate is imprecise given the few events, did not remain significant after adjustment for age, and should be regarded as hypothesis-generating.

By contrast, CAC was strongly and independently associated with subsequent cardiac testing utilization: women with calcium underwent nearly three times the rate of stress testing, coronary CT angiography, and invasive angiography, an association that persisted after adjustment for age. Because calcium was reported in only 5.4% of scans, this association is unlikely to be an artifact of clinicians acting on the calcium score itself; rather, CAC appears to mark a phenotype destined for greater cardiovascular evaluation. Recognizing calcium at baseline could help rationalize that testing, for example by informing the pre-test probability of downstream studies and the selection among them.

## Limitations

Our study has several limitations. It is retrospective and single-center, and the association between CAC and downstream testing may be influenced by unmeasured indications for that testing. Indications for downstream testing and symptom status at the time of testing were not available, so we cannot determine whether these evaluations were prompted by ischemic symptoms or by other, non-ischemic indications. The reclassification analysis reflects guideline eligibility rather than realized changes in prescribing or outcomes, and the prevention-eligible subgroup was modest in size. Eligibility was assessed with the 2019 ACC/AHA thresholds and the Pooled Cohort Equations in force during the study; the 2026 ACC/AHA dyslipidemia guideline, which adopts the PREVENT equations and reframes CAC around LDL-cholesterol goals, may alter the specific eligibility counts, although it continues to endorse CAC- guided statin allocation in this population.^6^ Follow-up for the primary outcome relied on administrative censoring, which may overestimate follow-up for patients lost to care; per-patient last-contact data and extended follow-up would refine the time-to-event analyses. Inter- and intra-reader reliability of the semi-automatic scoring were not available for this analysis and should be reported, particularly given evidence that Agatston scores derived from non-gated PET/CT may differ from gated measurement.^16^ CT parameters were those of routine oncologic PET/CT rather than a dedicated calcium protocol and varied across scanners and over time: tube voltage ranged from 100 to 140 kVp rather than the 120 kVp at which the 130-HU Agatston threshold is defined, and reconstructed slice thickness (3.0–4.0 mm) exceeded the 2.5–3.0 mm used for dedicated scoring. Because higher tube voltage and thicker sections tend to reduce measured calcium whereas lower tube voltage tends to increase it, absolute Agatston scores and derived percentiles should be regarded as approximations, and the direction and magnitude of any bias may differ by scanner. A complete lipid profile was unavailable for 37% of patients, so ASCVD and MESA risk scores could be calculated in 63%, and analyses involving them are complete-case; the frequent unavailability of complete risk-factor profiles further underscores the appeal of a marker derived from imaging already performed, requiring no additional testing. Ascertained cardiac events were restricted to atherosclerotic endpoints. Echocardiographic surveillance data were not available, so cancer therapy–related cardiac dysfunction, including asymptomatic reductions in left ventricular ejection fraction and any consequent interruption or modification of cancer therapy, was not ascertained; this study therefore cannot address whether CAC predicts cardiotoxicity. Details of cancer treatment, including anthracycline, HER-2–targeted, and radiotherapy exposure, were not abstracted and could not be examined as covariates or effect modifiers. Finally, events occurring outside our health system may have been incompletely captured.

## Conclusions

Coronary artery calcium can be feasibly and quantitatively scored from low-dose CT during staging ^18^F-FDG PET/CT in women with breast cancer, and doing so would frequently reclassify guideline-based statin eligibility in both directions, at no incremental cost or radiation exposure, yet calcium is rarely reported in current practice.

Standardized reporting of CAC on these already-acquired scans could convert a missed finding into actionable cardiovascular prevention at the earliest point in the cancer care pathway. A prospective study of whether such reporting changes prescribing and improves outcomes is warranted.

## Clinical Perspective

### What Is New?

- This is the first study in an all-female breast-cancer cohort to show that a discrete, percentile-capable Agatston coronary artery calcium score can be derived semi-automatically from the low-dose CT of a staging ^18^F-FDG PET/CT obtained before cancer therapy, providing the level of quantification needed to apply the full set of guideline statin thresholds, including the 75th-percentile trigger, from imaging these women already undergo.
- Applying guideline thresholds to this score would reclassify statin eligibility in nearly two-thirds of prevention-eligible women, initiating therapy in the under-treated and supporting de-prescribing where calcium is absent.

### What Are the Clinical Implications?

- Standardized reporting of coronary calcium already visible on staging PET/CT could enable actionable cardiovascular prevention at the earliest point in breast-cancer care, at no additional cost or radiation exposure.
- A prospective study is needed to determine whether such reporting changes prescribing and improves clinical outcomes.

## Acknowledgments

None.

## Non-standard Abbreviations and Acronyms

ASCVD: atherosclerotic cardiovascular disease
AU: Agatston units
CAC: coronary artery calcium
CAC-A: coronary artery calcium absent
CAC-P: coronary artery calcium present
CACS: coronary artery calcium score
CCTA: coronary computed tomography angiography
CVD: cardiovascular disease
MACE: major adverse cardiac events
MESA: Multi-Ethnic Study of Atherosclerosis
PET/CT: positron emission tomography/computed tomography

## References

1. Mehta LS, Watson KE, Barac A, et al. Cardiovascular Disease and Breast Cancer: Where These Entities Intersect: A Scientific Statement From the American Heart Association. Circulation. 2018;137(8):e30–e66.

2. Patnaik JL, Byers T, DiGuiseppi C, Dabelea D, Denberg TD. Cardiovascular disease competes with breast cancer as the leading cause of death for older females diagnosed with breast cancer: a retrospective cohort study. Breast Cancer Res. 2011;13(3):R64.

3. Lenihan DJ, Cardinale DM. Late cardiac effects of cancer treatment. J Clin Oncol. 2012;30(30):3657–3664.

4. Darby SC, McGale P, Taylor CW, Peto R. Long-term mortality from heart disease and lung cancer after radiotherapy for early breast cancer: prospective cohort study of about 300,000 women in US SEER cancer registries. Lancet Oncol. 2005;6(8):557–565.

5. Koczo A, Brickshawana A, Zhu J, et al. Sex-Based Utilization of Guideline-Recommended Statin Therapy and Cardiovascular Disease Outcomes: Data from a Multisite Healthcare Network Primary Prevention Cohort. Am J Prev Cardiol. 2024;18:100672.

6. Blumenthal RS, Morris PB, Gaudino M, et al. 2026 ACC/AHA/AACVPR/ABC/ACPM/ADA/AGS/APhA/ASPC/NLA/PCNA Guideline on the Management of Dyslipidemia: A Report of the American College of Cardiology/American Heart Association Joint Committee on Clinical Practice Guidelines. J Am Coll Cardiol. Published online March 13, 2026. doi:10.1016/j.jacc.2025.11.016.

7. Arnett DK, Blumenthal RS, Albert MA, et al. 2019 ACC/AHA Guideline on the Primary Prevention of Cardiovascular Disease. Circulation. 2019;140(11):e596–e646.

8. Nasir K. Message for 2018 Cholesterol Management Guidelines Update: Time to Accept the Power of Zero. J Am Coll Cardiol. 2018;72(25):3243–3245.

9. Lopez-Mattei J, Yang EH, Baldassarre LA, et al. Cardiac computed tomographic imaging in cardio-oncology: An expert consensus document of the Society of Cardiovascular Computed Tomography (SCCT). J Cardiovasc Comput Tomogr. 2023;17(1):66–83.

10. Lyon AR, Lopez-Fernandez T, Couch LS, et al. 2022 ESC Guidelines on cardiooncology. Eur Heart J. 2022;43(41):4229–4361.

11. Raisi-Estabragh Z, Murphy AC, Ramalingam S, et al. Cardiovascular Considerations Before Cancer Therapy: Gaps in Evidence and JACC: CardioOncology Expert Panel Recommendations. JACC CardioOncol. 2024;6(5):631–654.

12. Sandhu AT, Rodriguez F, Ngo S, et al. Incidental Coronary Artery Calcium: Opportunistic Screening of Previous Nongated Chest Computed Tomography Scans to Improve Statin Rates (NOTIFY-1 Project). Circulation. 2023;147(9):703–714.

13. Foraker R, Sperling L, Bratzke L, Budoff M, Leppert M, Razavi AC, et al. Opportunistic Detection of Coronary Artery Calcium on Noncardiac Chest Computed Tomography: A Scientific Statement From the American Heart Association. Circulation. 2025;152:e391–e401.

14. Osborne-Grinter M, Ali A, Williams MC. Prevalence and clinical implications of coronary artery calcium scoring on non-gated thoracic computed tomography: a systematic review and meta-analysis. Eur Radiol. 2024;34(7):4459–4474.

15. Mais HE, Kay R, Almubarak H, et al. Prognostic importance of coincidental coronary artery calcification on FDG-PET/CT oncology studies. Int J Cardiovasc Imaging. 2021;37(4):1479–1488.

16. Pak S, Son HJ, Kim D, et al. Standard Visual and Ordinal Coronary Calcium Scoring on PET/CT: Agreement with Agatston Scoring and Prognostic Implications. Diagnostics (Basel). 2025;15(23):2969.

17. Morf C, Sartoretti T, Gennari AG, et al. Diagnostic Value of Fully Automated Artificial Intelligence Powered Coronary Artery Calcium Scoring from 18F-FDG PET/CT. Diagnostics (Basel). 2022;12(8):1876.

18. Mazet R, Torossian N, Wanneveich M, et al. Calcium scoring during 18F-FDG PET/CT in cancer indications: Improving cardiovascular risk stratification and prevention. PLoS One. 2025;20(10):e0335249.

19. Phillips WJ, Johnson C, Law A, et al. Comparison of Framingham risk score and chest-CT identified coronary artery calcification in breast cancer patients to predict cardiovascular events. Int J Cardiol. 2019;289:138–143.

20. Gal R, van Velzen SGM, Hooning MJ, et al. Identification of Risk of Cardiovascular Disease by Automatic Quantification of Coronary Artery Calcifications on Radiotherapy Planning CT Scans in Patients With Breast Cancer. JAMA Oncol. 2021;7(7):1024–1032.

21. Goff DC Jr, Lloyd-Jones DM, Bennett G, et al. 2013 ACC/AHA guideline on the assessment of cardiovascular risk. Circulation. 2014;129(25 Suppl 2):S49–S73.

22. McClelland RL, Nasir K, Budoff M, et al. Arterial Age as a Function of Coronary Artery Calcium (from the Multi-Ethnic Study of Atherosclerosis [MESA]). Am J Cardiol. 2009;103(1):59–63.

23. McClelland RL, Jorgensen NW, Budoff M, Blaha MJ, Post WS, Kronmal RA, et al. 10-Year Coronary Heart Disease Risk Prediction Using Coronary Artery Calcium and Traditional Risk Factors: Derivation in the MESA With Validation in the HNR and DHS. J Am Coll Cardiol. 2015;66(15):1643–1653.

24. Agatston AS, Janowitz WR, Hildner FJ, Zusmer NR, Viamonte M Jr, Detrano R. Quantification of coronary artery calcium using ultrafast computed tomography. J Am Coll Cardiol. 1990;15(4):827–832.

25. McClelland RL, Chung H, Detrano R, Post W, Kronmal RA. Distribution of coronary artery calcium by race, gender, and age: results from the Multi-Ethnic Study of Atherosclerosis (MESA). Circulation. 2006;113(1):30–37.

26. Thygesen K, Alpert JS, Jaffe AS, et al. Fourth Universal Definition of Myocardial Infarction (2018). J Am Coll Cardiol. 2018;72(18):2231–2264.

27. Messenger B, Li D, Nasir K, Carr JJ, Blankstein R, Budoff MJ. Coronary calcium scans and radiation exposure in the multi-ethnic study of atherosclerosis. Int J Cardiovasc Imaging. 2016;32(3):525–529.

28. Roberts ET, Horne A, Martin SS, et al. Cost-effectiveness of coronary artery calcium testing for coronary heart and cardiovascular disease risk prediction to guide statin allocation: the Multi-Ethnic Study of Atherosclerosis (MESA). PLoS One. 2015;10(3):e0116377.

29. Newman CB, Preiss D, Tobert JA, et al. Statin Safety and Associated Adverse Events: A Scientific Statement From the American Heart Association. Arterioscler Thromb Vasc Biol. 2019;39(2):e38–e81.

30. Otte JL, Skaar TC, Wu J, et al. Medication use in breast cancer survivors compared to midlife women. Support Care Cancer. 2013;21(7):1827–1833.

31. Iliescu CA, Grines CL, Herrmann J, et al. SCAI Expert consensus statement: Evaluation, management, and special considerations of cardio-oncology patients in the cardiac catheterization laboratory. Catheter Cardiovasc Interv. 2016;87(5):E202–E223.

32. Dzaye O, Al Rifai M, Dardari Z, et al. Coronary Artery Calcium as a Synergistic Tool for the Age- and Sex-Specific Risk of Cardiovascular and Cancer Mortality: The Coronary Artery Calcium Consortium. J Am Heart Assoc. 2020;9(8):e015306.

